# Chest X-Ray as a critical screening tool for Household Contacts of TB: Lessons from Three Years of Programmatic Data in India

**DOI:** 10.64898/2026.06.16.26355768

**Authors:** Ridhima Sodhi, Pranati Das, Ashwani Khanna, Veena Dhawan, Radha Taralekar, Harkesh Dabas, Shamim Mannan, Manoj Singh

**Affiliations:** William J Clinton Foundation, New Delhi, India; Central TB Division, Ministry of Health & Family Welfare, Government of India

## Abstract

**Introduction:** Household contacts (HHCs) of pulmonary TB patients remain at high risk for TB infection and disease progression, yet many remain asymptomatic and are missed by symptom-screening pathways. While India expanded its TB preventative guidelines to include all HHCs in 2021, chest X-ray (CXR) screening continues to be used selectively, representing a missed opportunity in early case detection.

**Methods:** The analysis uses programmatic data from Project JEET 2.0 (Joint Effort for Elimination of Tuberculosis), implemented by the William J. Clinton Foundation in India, between October 2021 and March 2024. Eligible HHCs (>=5 years) were offered CXR screening as part of TB preventive therapy (TPT) evaluation. Descriptive and multivariable analyses examined predictors of CXR uptake and TB yield. A two-stage logistic regression model estimated potential TB yield under universal CXR coverage. Model performance was evaluated using the area under the curve (AUC), and bootstrap simulations generated counterfactual estimates of missed TB cases.

**Results:** Among 1,034,621 HHCs, 1.02% individuals were found positive for TB, which includes 7,786 HHCs who were on TB treatment already, while an additional 2,812 were identified during pre-TPT evaluation. Among eligible HHCs (n = 1,026,835), 70% were screened with CXR, of which 2.4% had suggestive TB findings. Of these, 79% went for further TB assessment. Symptomatic HHCs were more likely to be CXR screened (84% vs 69%) and assessed for TB, yet two-thirds of all detected TB cases were asymptomatic. It is estimated that universal CXR coverage and TB testing for suggestive cases can increase TB detection by at least 87%.

**Conclusion:** The study provides a scalable approach to expand CXR coverage through public-private partnerships, enabling early TB detection among HHCs, especially among asymptomatic contacts. Future implementations will benefit from integrating AI-enabled reading, along with systematic follow up for those with suggestive findings.

## Introduction

India carries 25% of the global TB burden, of which a large majority of individuals remain undetected by health systems^1–3^; national estimates suggest that two TB patients are missed for every one diagnosed^1^. Household contacts (HHCs) of TB patients are at a particularly high risk, a larger majority is infected with TB infection (TBI), which refers to the presence of *Mycobacterium tuberculosis* in an individual, albeit in a dormant state^1^. The susceptibility of progressing to active TB disease among HHCs varies, and is higher among those with existing comorbidities, malnourishment, or those that live or/and work in poorly ventilated conditions^4^. India recently expanded its TB preventive treatment (TPT) programme to ensure systematic evaluation of all close contacts, going beyond mandating the treatment for under-5 children and people living with HIV (PLHIV)^2,4,5^.

Initiating TBI treatment among HHCs, however, needs to be pre-empted by ruling out of active TB disease; and should ideally succeed TBI determination^5,6^. Identifying TBI carries its own challenges; the widely used tuberculin skin test (TST) has limited accuracy in India, and interferon-gamma release assays (IGRAs) remain cost-prohibitive^7,8^. Additionally, while national TPT guidelines recommend using Chest X-ray (CXR) as a pre-screening measure to rule out TB, the said measure is not mandated, and often triggered only post a positive symptomatic assessment^5^. This particularly risks asymptomatic HHCs, who form a large reservoir for both TBI and active TB^9,10^.

Recognizing this, the William J. Clinton Foundation, while implementing the TPT program in India, incorporated CXR screening for all HHCs prior to TPT initiation. This study aims to estimate the impact of such screening on TB yield among HHCs, particularly among asymptomatic individuals. Drawing upon programmatic data from 71 districts across 11 states and union territories in India, it provides an overview of how such interventions can be integrated into broader public health strategies.

## Methods

### Study Setting - Latent TB services under Project JEET 2.0

Between October 2021 and March 2024, the Global Fund supported the expansion of TPT program in India across 207 districts in 23 states and union territories, under Project JEET 2.0 (Joint Effort for Elimination of Tuberculosis). Implemented in collaboration with the National TB Elimination Programme (NTEP), JEET 2.0 was led by three agencies: FIND (5 states, 28 districts), The Union (7 states, 108 districts), and the William J. Clinton Foundation (WJCF) (11 states, 71 districts). This study draws on programmatic data from the WJCF component implemented across Bihar, Delhi, Gujarat, Haryana, Kashmir, Kerala, Ladakh, Rajasthan, Tamil Nadu, Uttarakhand, and Uttar Pradesh.

Project beneficiaries included HHCs of pulmonary drug-sensitive TB (PTB) patients aged >=5 years, as TPT for children aged <5 years has been provisioned and indicated for all without TBI testing under NTEP guidelines ^5^. Other exclusions include people living with HIV, and individuals already on TB treatment or TPT. Pre-TPT evaluation was offered to all beneficiaries, and included: a) WHO recommended four-symptom screen (W4SS) ^9^, and b) CXR screening. CXR was offered free through public X-ray facilities; however, only 86% of the 758 treatment units (TUs) had a functional public-sector unit, often affected by breakdowns, staff shortages, restricted hours, and long waiting times. To ensure access, private laboratories were empanelled in 91% of TUs through a voucher-based payment model, expanding CXR availability to 98% of TUs (744/758). Despite this, uptake remained limited in rural areas, where distance and low perceived risk among asymptomatic contacts reduced participation.

### Study Design

This retrospective programmatic analysis evaluated the impact of CXR screening on TB detection among HHCs of PTB patients. Descriptive analyses summarised cohort characteristics, and multivariable methods were applied to verify observed trends. A two-stage logistic regression model estimated TB yield under universal CXR coverage. Bootstrapping was used to generate confidence intervals and assess model stability across heterogeneous programmatic contexts.

### Patient and public involvement

This study was based on secondary analysis of routinely collected, anonymised programmatic data, and there was no direct contact with patients. As such, it was not appropriate or feasible to involve patients or members of the public in the research process.

### Study population

The cohort comprised HHCs of 304,107 PTB patients notified in Nikshay, India’s national TB database. Home visits were conducted for contact listing, symptom screening, counselling, and linkage to services. HHCs were defined as individuals sharing an enclosed living space with the index patient, following national TPT guidelines ^5^. Symptomatic contacts were defined as those reporting any one or more of the WHO four-symptom screen (W4SS) symptoms: cough ≥2 weeks, fever, weight loss, night sweats; with haemoptysis or coughing up of blood being noted as the fifth symptom ^5,9^. A suggestive CXR referred to radiographic abnormalities consistent with TB, assessed by radiologists or trained medical officers. TB diagnosis was based on microbiological confirmation or clinical diagnosis by a qualified medical officer.

### Data Sources

Three data sources were used in this study. The primary dataset used includes the LTBI 360 the JEET2.0 Project Management Information System (MIS), which contained information on socio-demographic characteristics (age, date of birth, gender, marital status) and clinical details (visit, screening, and testing dates; symptom profile; risk factors; TB treatment history; TPT initiation and completion dates; and regimen type). The second dataset referred to those with TB-suggestive CXR findings, including information on subsequent diagnostic testing and TB diagnosis. Nikshay, or the government’s national data portal for TB management was used to supplement treatment outcome data recorded by healthcare workers. The datasets were linked using the unique identification key assigned to each household contact.

### Analysis

A combination of descriptive and regression analyses were used. Population characteristics were summarized using descriptive statistics, and group differences were assessed using Kruskal-Wallis rank-sum tests for continuous variables and chi-square tests for categorical or dichotomous variables. Logistic regression models were used to estimate adjusted associations of CXR uptake and TPT initiation with covariates. Results are presented as adjusted odds ratios (aOR) with robust 95% confidence intervals (CI), with standard errors clustered at the state level. Covariates included demographic characteristics (age, gender), microbiological confirmation status of the index patient, diagnostic evaluation for ruling out TB (CXR screening), diagnostic evaluation for TBI (IGRA, TST), occupational status, symptom profile, state, and diagnosing quarter of index patient. Occupational status was missing for 27% of observations; to retain sample size, missing values were included as a separate “unknown” category in regression models. To assess the potential gains from expanding CXR coverage to all household contacts (HHCs), a two-stage logistic model was fitted. Model discrimination was evaluated using the area under the receiver operating characteristic curve (AUC), and 95% confidence intervals were derived using nonparametric bootstrapping.

### Robustness Checks

Several robustness checks were conducted to assess the stability of the findings. Alternative model specifications included (i) excluding observations with missing occupational status (i.e., those classified as “unknown”), and (ii) excluding contacts from the state of Kerala, where additional state-sponsored CXR screening was implemented but not captured in the available data. Findings remained consistent across these specifications. In addition, nonparametric bootstrapping with 200 resamples was used to generate robust confidence intervals for the two-stage logistic model estimating the impact of universal screening and TB diagnostics.

### Data Selection (Inclusion & Exclusion criteria)

The initial dataset included 1,224,464 HHCs. Sequential exclusions were applied for non-screening or lack of consent, age under five years, incomplete or miscoded CXR status, or referral due to comorbidities. The final analytic cohort included 1,026,835 HHCs, of whom 7,779 (0.76%) were already on TB treatment, leaving 1,026,835 HHCs eligible for TPT under JEET 2.0 program, and our analysis (Table S1).

### Software

Analyses were conducted in R (2025.09.1+401) using standard statistical packages for data management (dplyr, tidyr) and modelling (glm, sandwich).

### Ethical Considerations

A study protocol was approved by the Monk Prayogshala Institutional Review Board and Ethics Review Committee on 26/08/2025, which granted a waiver of informed consent as the analysis relied on anonymised and de-identified programmatic data. The data were accessed for research purposes on 29/08/2025.

## Results

### Demographic Summary

Available data included information on 1,034,621 HHCs, of which 0.7% were already on TB treatment (Figure 1), hence not eligible for TPT evaluation. Table 1 presents summary statistics for the 1,026,835 HHCs who were screened; of which 52% were females, and ~0.02% were transgender. Majority of HHCs were from Uttar Pradesh (42%), distantly followed by Bihar (16%), Rajasthan (11%), and Gujarat (10%). HHCs screened with CXR made 70% of the cohort, of which 2.3% received a suggestive CXR reading. Among those with a suggestive TB reading in CXR, 79% went in for further TB assessment; 21% (2,812) were diagnosed with TB.

**Table 1.**
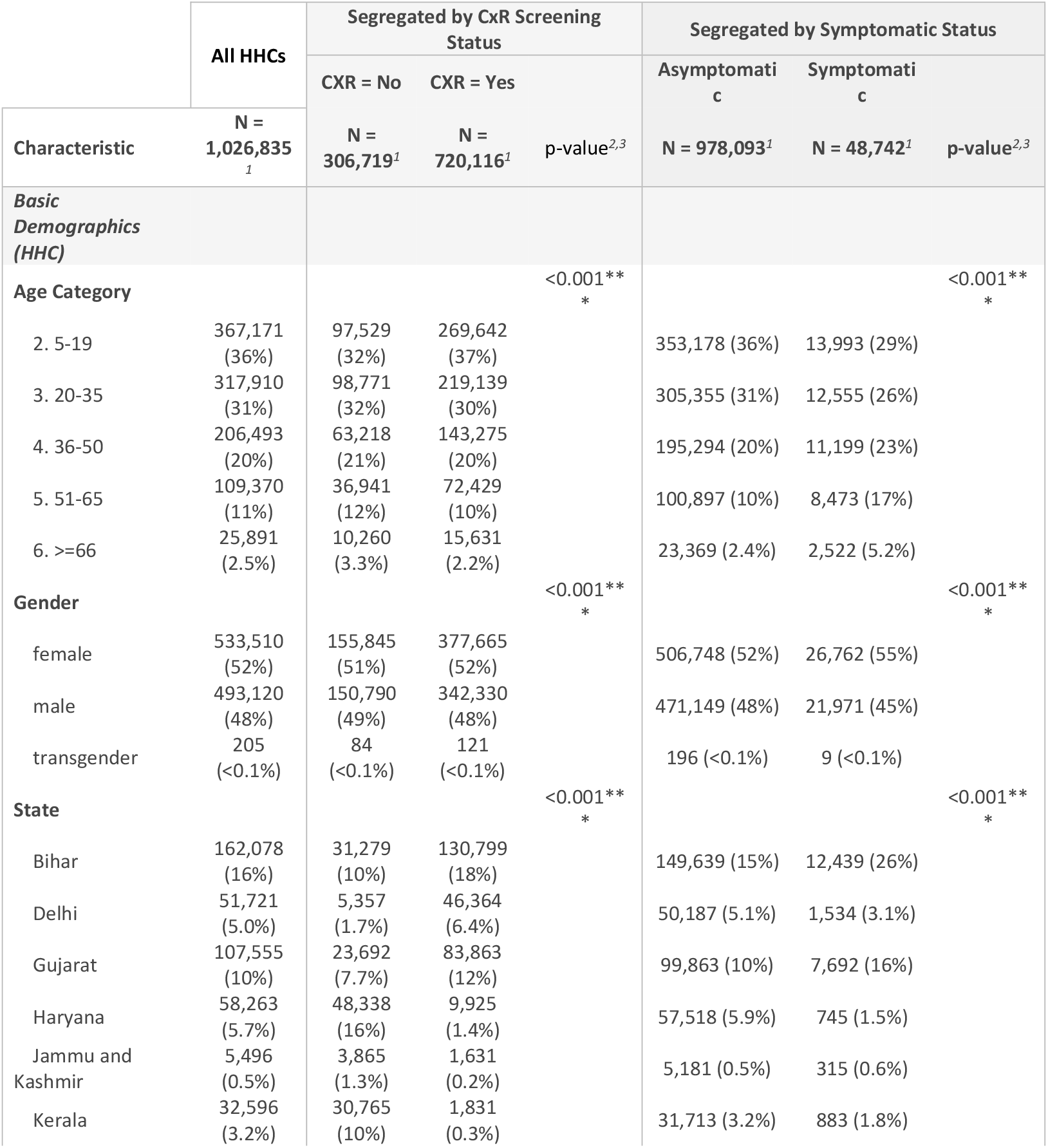
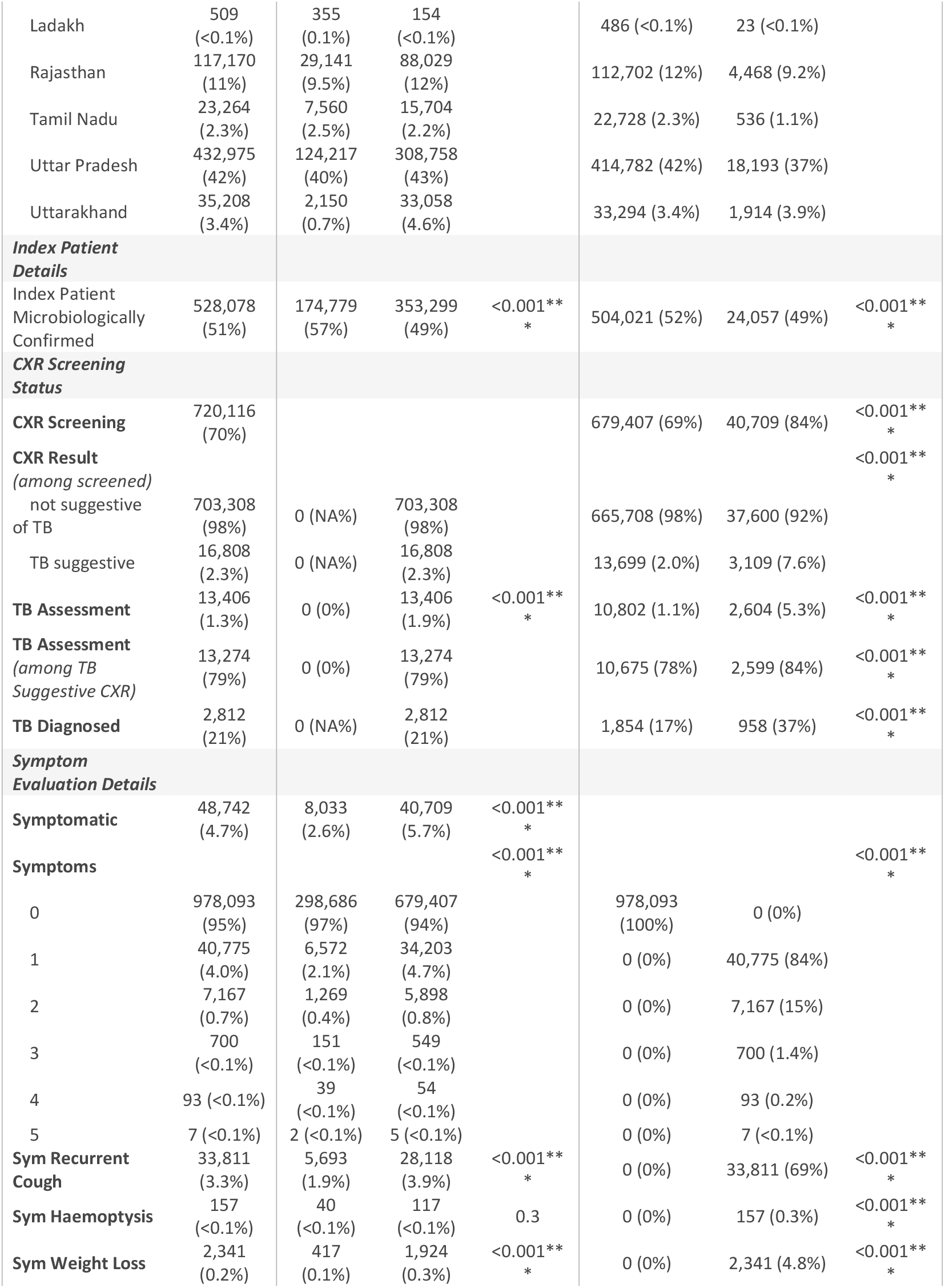
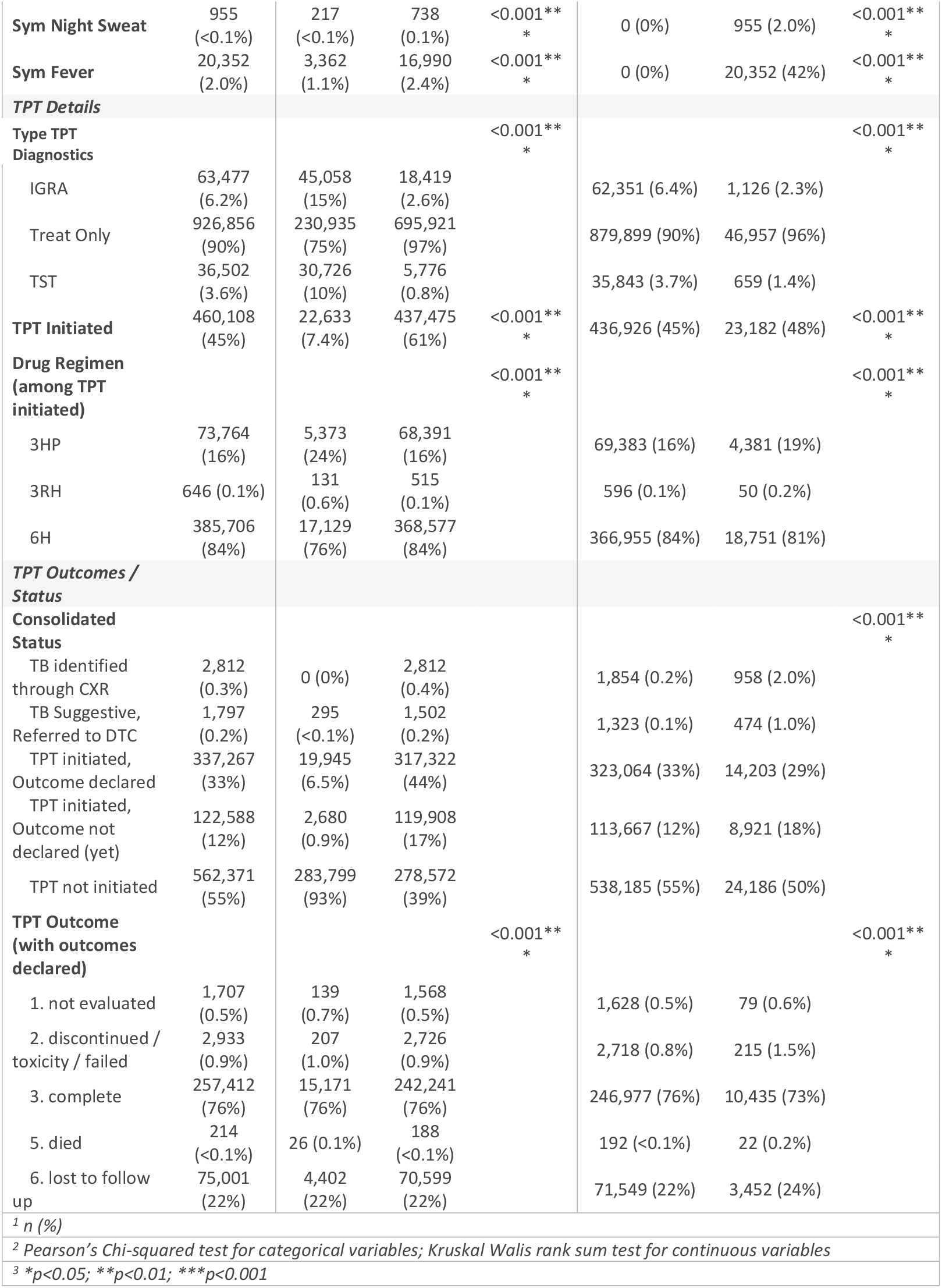
Summary Statistics – Overall, segregated by CXR Screening Status, and Symptomatic Status; N = 1,026,835.

**Figure 1.**
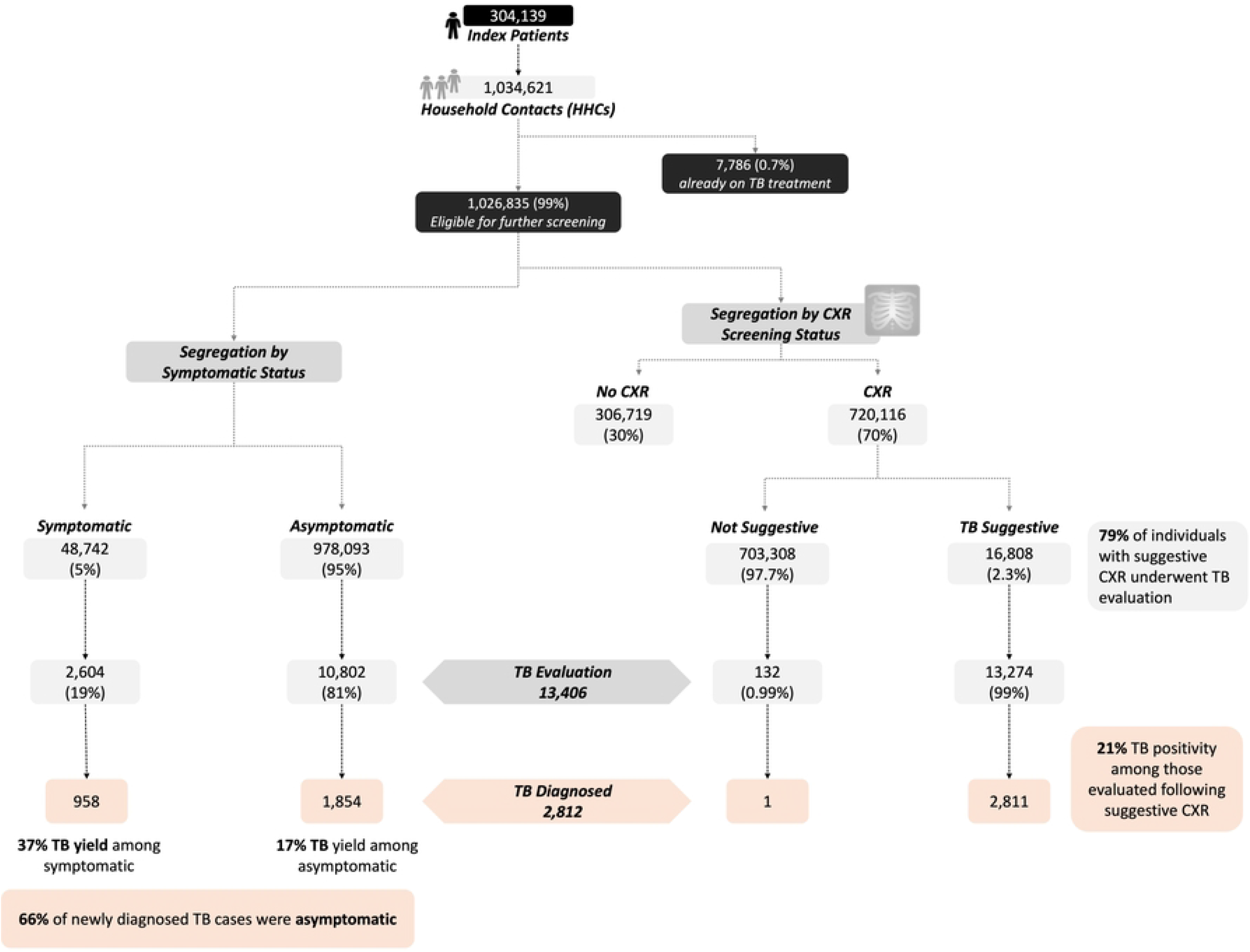
Observed TPT screening and TB assessment cascade; N = 1,034,621 HHCs.

### Factors associated with CxR screening

Overall, 70% of household contacts (HHCs) underwent CXR screening (Table 1), with substantial regional variation (Table S2). Screening coverage exceeded 90% in Delhi and Uttarakhand but was considerably lower in Haryana (17%) and Kerala (6%). In Kerala, CXR uptake likely reflects under-ascertainment, as HHCs in the state are known to have received CXR from public facilities, data for which has not been made available; CXR uptake here only refers to what is provided by the JEET 2.0 program. Screening coverage increased over time, from 44% in 2021 Q4 to 71% in 2024 Q1 (Table S3).

Screening uptake was strongly associated with symptomatic status (aOR 1.73; 95% CI: 1.63-1.84) (Model 1, Table S4), with 84% of symptomatic HHCs screened compared to 69% of asymptomatic individuals (Table 1C). Older adults (≥66 years) were less likely to be screened (aOR 0.69; 95% CI: 0.60-0.80), with 60% coverage compared to 73% among those aged 5-19 years (Table S5). HHCs engaged in agriculture, livestock, and fisheries had lower screening uptake (62%) (Table S6), although this association was not statistically significant. Screening did not differ by whether the index patient was microbiologically confirmed (aOR 1.04; 95% CI 0.96-1.13).

### CxR screening & TPT Initiation

TPT initiation was substantially higher among those screened with CXR (61%) compared with those not screened (7.4%) (Table 1B); reflected also in the logistic model (aOR 64.14; 95% CI: 29.12-141.27) (Model 1, Table S7). Overall, majority of TPT initiations occurred among individuals who had undergone CXR screening (437,435/460,068; 95%).

Microbiological confirmation of index patients was associated with higher TPT initiations (aOR: 1.15; 95% CI: 1.06, 1.24); although this resulted in only a 2 percentage point difference between the two groups (46% vs 44%, Table S8). Symptoms were negatively associated, but the association was only mildly significant (aOR: 0.86; 95% CI: 0.76, 0.97). This is consistent with higher symptoms also corresponding with suggestive TB readings in CXR, and consequently TB referrals (Tables S9, S10).

### TB Assessments & Diagnostic Methods

79% (13,406) of those with a suggestive reading went for further TB assessment (Table 1B), where the diagnosis was significantly affected by symptomatic status (Table S9). Logistically, every additional symptom increased the odds of TB assessment by 24% (aOR: 1.24; 95% CI: 1.09, 1.42) (Model 1, Table S10), resulting in a 6 percentage point difference between TB assessments among the symptomatic and asymptomatic groups (84% vs 78%) (Table 1C). Bacteriological confirmation of TB among index patients did not impact TB assessment or TB positivity among HHCs (21% in each cohort among those who were assessed) (Table S8). Diagnostic methods varied – only 17% got a NAAT, and 43% relied on clinical assessment (Table 2). TB positivity rates differed significantly (p<0.001), and were high for NAAT (43%), and low for clinical (15.3%) and microscopy (16%) (Table S11).

**Table 2.**
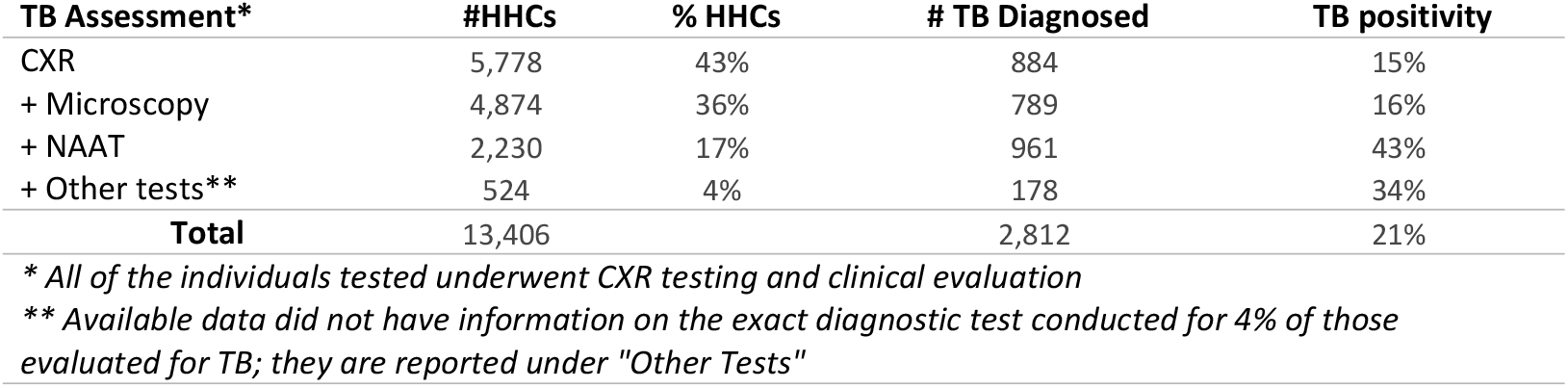
TB Diagnostic Methods (N = 13,406)

### TB Prevalence and Symptoms

*Identified* TB yield among all screened HHCs was 1.3%, considering the 7,786 HHCs who were already on TB treatment, and the 2,812 individuals found through screening and TB assessments (Figure 1). While symptomatic were more likely to be TB diagnosed (37% vs 17%, p<0.001) (Table 1C), asymptomatic make up 81% of assessed, and 66% of those with a positive TB diagnosis (Figure 1). This is attributable to the fact that asymptomatic make up 95% of the cohort, and asymptomatic tested (n = 10,802) are more than four times that of symptomatic tested (n = 2,604).

### Projected Impact of Universal Screening

Among those not already on TB treatment at the time of screening, observed TB yield was 0.28% (2,812/1,026,835). The actual TB yield among this cohort is expected to be higher, on at least three accounts. First, 30% (n=306,719) of HHCs did not undergo CXR screening, majority of which were also not initiated on TPT (Table S12). Second, 21% (3,534/16,808) of individuals with suggestive CXR findings did not undergo TB assessment: 8% of these started TPT, and another 14% were referred for TB assessment, albeit further information is not available. It is hence, plausible that additional TB cases existed among these HHCs (n = 310,253). A two-stage logistic mode was fitted to estimate impact of universal screening. The first model estimated the probability of a suggestive CXR finding among those screened (Table S13, Figure 2A), while the second estimated the probability of TB positivity among individuals with suggestive CXRs who were assessed (Table S14, Figure 2B); described also as below:

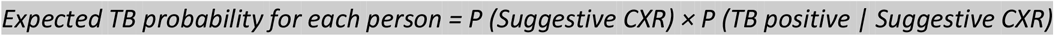

A third reason, which while not factored in this projection, posits our TB yield as underestimated is that only 17% relied on NAAT testing. This was not modelled in our analysis, as we do not have sufficient information to estimate by how much universal NAAT testing would affect TB diagnosis, even though the data suggests that the effect will be positive.

**Figure 2.**
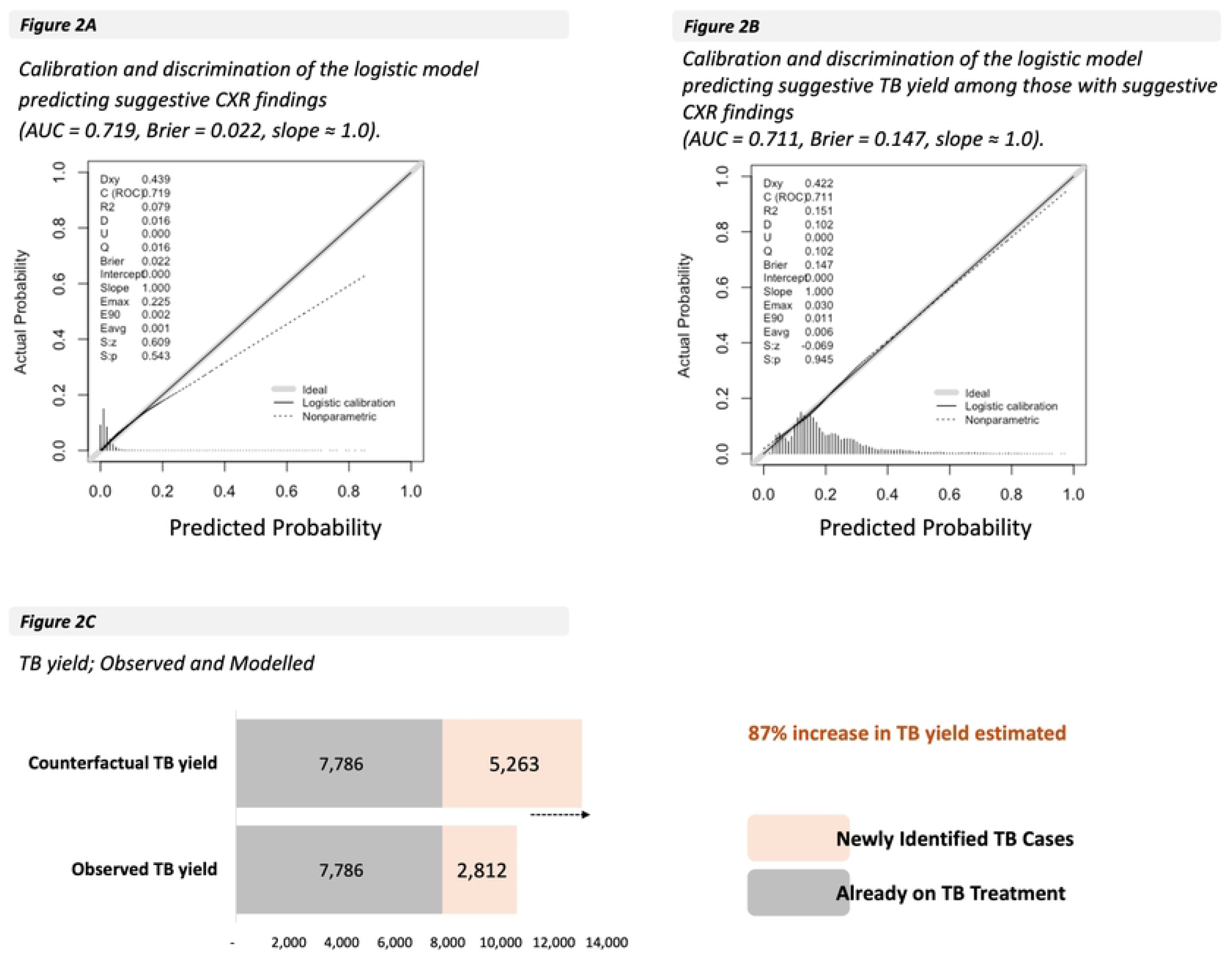
Two Stage Counterfactual Model; Calibration and Discrimination of the Logistic Models & Modelled TB Yield.

The two models had areas under the curve (AUC) of 0.719 and 0.711, indicating that in approximately 71% of cases, a person with TB was correctly ranked as higher risk than a person without TB, indicating adequate separation between TB and non-TB cases. The estimated coefficients were used to simulate TB diagnosis in the case of universal CXR screening and TB assessments for all those with suggestive readings. Predicted TB yield was estimated as 0.51% (95% CI: 0.511%-0.515%), higher than the observed prevalence of 0.273% (95% CI: 0.264 – 0.284). Estimated TB cases identified as per the model are 5,263, representing an 87% increase in case detection (relative to 2,812 identified prior) (Figure 2).

## Discussion

The study represents the largest evaluation of CXR screening among HHCs (>=5 years) of drug-sensitive PTB patients. The findings reinforce the critical role of CXR-based screening in reducing the TB detection gap, particularly among asymptomatic HHCs. The findings align with emerging global evidence on subclinical and early TB among contacts, albeit in a much larger, geographically diverse setting^11–13^.

### Asymptomatic Contacts

India continues to face a considerable gap between estimated and reported TB incidence^1–3^, driven in part by reliance on symptom-based assessment alone^9,14,15^. In our cohort, asymptomatic HHCs constituted 95% of all HHCs and 66% of those diagnosed with TB. Including asymptomatic screening, hence, accounted for a 200% increase in TB detection (958 −> 2,812); substantially higher than the 75% increase reported from a similar contact investigation program in Chennai^13^. Studies from Peru and Indonesia have also demonstrated abnormal CXR findings to be strongly predictive of TB progression among asymptomatic contacts^11,12^. Without CXR, many of these individuals might have been misclassified as having TBI, started inappropriately on TPT, or missed entirely.

In our cohort, 21% (n = 3,534) of those with TB suggestive CXR readings weren’t followed up, of which 86% (n = 3,024 were asymptomatic to begin with; echoing concerns from other settings that symptom and CXR algorithms alone still miss or delay detection of a substantial proportion of asymptomatic TB^11^. Screening asymptomatic contacts, however, is operationally challenging. First, a lack of symptoms is often linked to lower risk perception; and second, there is a general lack of awareness about TPT, among both patients and doctors^16–18^. Behaviour and psychosocial support may help here; a recent systematic review of psychosocial interventions found that tailored education, survivor testimonials and community-based, multidisciplinary support can increase TPT initiation by 10–52%. Cost and feasibility are additional concerns, given the large size of the asymptomatic contact pool. However, evidence suggests that CXR-based screening is cost-favourable in high-burden settings when integrated within existing contact-tracing systems^19,20^.

### Testing HHCs of all Index Patients, irrespective of diagnostic method

The study also contributes to the growing evidence of TBI & TB positivity among contacts of clinically diagnosed TB patients^17,21,22^; even when conventional strategies continue to target contacts of bacteriologically confirmed PTB^13,21^. In this highly representative cohort (of India), the clinically and microbiologically diagnosed groups were largely evenly split; with no significant difference in underlying TB positivity; highlighting the significant gap in traditional contact strategies. Emerging literature emphasizes the need to include this group in contact tracing, alongside other typically excluded groups such as those with extrapulmonary TB^21^.

### Diagnostic Methods & TB Positivity among HHCs

While recommended by NTEP for many benefits including higher sensitivity, drug-sensitivity testing, and rapid turnaround^23–26^, only 17% of those getting assessed for TB in this cohort underwent such testing. Availability of NAAT testing is constrained by challenges such as high cost^26–28^, and unavailability of cartridges^29^; likely influencing decisions at both the patient and program level. CXR screening can work as a triage here; recent cluster-randomized data from Benin and Brazil show that strategies using targeted CXR or rapid molecular tests can achieve high TPT initiation at acceptable societal costs, while avoiding unnecessary investigations in truly low-risk contacts^30^. Rapid advances in NAAT technology, especially portable and point-of-care platforms^31,32^, alongside public-private partnerships can be pivotal to improve equitable access to both CXR and NAAT^13,26,33^. Programmatic experiences from India and other countries demonstrate the role of such approaches in increasing diagnostic reach, without overburdening the public sector^34–36^. The increasing availability of digital CXR and AI-based computer-aided detection (CAD) tools are likely to enhance equitable access, particularly in resource-limited or decentralized settings^37^.

### Missed HHCs – Identified Gaps

Merely provisioning CXR, however, does not ensure universal screening. Barriers including limited access to functional X-ray facilities, restricted operating hours, long waiting times, and travel costs were consistently reported across JEET 2.0 implementation sites, mirroring findings from other Indian and international TPT cascade studies^6,38–40^. Provider reluctance toward TPT is known to further compound these gaps, underscoring the role of building awareness and demand^38,41,42^. Follow-up after suggestive CXR findings remains an equally critical challenge: in our cohort, 21% of individuals with TB-suggestive readings did not proceed to TB assessment, of whom 86% were asymptomatic. Evidence from Malawi and Kenya corroborates this pattern; even when initial screening is prompt, losses accumulate between CXR and clinical decision-making, driven by both health system constraints and patient-level factors^40^. Timeliness analyses from Kenya further highlight that even when line-listing and initial screening are prompt, losses often occur between screening and clinical decisions on TB or TPT, driven by similar health system and patient-level constraints^43^. Our model estimates an 87% increase in TB diagnoses under universal CXR coverage and systematic diagnostic testing, and this figure is likely conservative. Suggestive CXR yields in our cohort (2.4%) were lower than those reported in other active case-finding studies^44^, where CXR-based strategies combined with broader sputum testing have identified substantially higher proportions of bacteriologically confirmed TB, including predominantly asymptomatic or pauci-bacillary disease^11,13,45^. Furthermore, only 17% of assessed HHCs received NAAT testing, which showed a positivity rate of 43% compared to 15-16% for clinical assessment or microscopy alone, suggesting that true TB burden among contacts is meaningfully higher than what programmatic data captures. Closing the gap between actual and identified TB cases will require strengthened referral linkages, mandatory follow-up protocols, and investment in diagnostic capacity, in both India, and other high-burden geographies.

## Conclusion

This study provides the largest real-world evidence to date on CXR-based screening within a TPT contact investigation program. With nearly two-thirds of TB-positive individuals being asymptomatic, the findings illustrate the dual risk of symptom-based triage: missed TB diagnoses and inappropriate TPT initiation among undetected TB-positive contacts. The study also demonstrates how empanelling private laboratories through a voucher-based payment model can enable universal CXR screening at scale, particularly where public-sector infrastructure remains unreliable. Building on this, integrating such models more stringently within NTEP, alongside mandatory follow-up for suggestive findings and wider NAAT adoption, has the potential to substantially narrow India’s TB detection gap. With global momentum building around TB elimination, systematic CXR screening for all household contacts, regardless of symptom status, should be treated not as an operational aspiration, but as a programmatic standard.

## Abbreviations

W4SS: WHO-recommended four-symptom screen
JEET: Joint Effort for Elimination of Tuberculosis
OLS: Ordinary least squares
CI: Confidence interval
PTB: Pulmonary Drug Sensitive aOR – Adjusted Odds Ratio
TB: Tuberculosis
NTEP: National Tuberculosis Elimination Program
NSP: National Strategic Plan for Elimination of Tuberculosis

## Supplementary Information

S1 Appendix: Supplementary Results

S2 Appendix: Gazette Notification; March 2018; TB a notifiable disease

## Limitations

The study was conducted within the programmatic framework of JEET 2.0 operations, and certain limitations are acknowledged. **First**, this analysis is based on routine programmatic data and is therefore subject to potential reporting and classification errors. Information on behavioural and structural determinants, such as stigma, distance to facility, household income, or health-seeking behaviour, was not available, limiting the ability to assess their influence on screening participation or diagnostic completion. **Second**, the logistic model was developed using data from individuals who underwent CXR screening and subsequently applied to the entire cohort to estimate potential missed TB cases. While this approach leverages a large, multi-state dataset to enhance generalizability, residual bias arising from differences between screened and unscreened contacts cannot be completely ruled out. **Third**, we have two reasons which suggest that our TB prevalence estimates are likely to be conservative, and the true burden of TB among household contacts may be higher than reported. This includes a) we did not model a diagnostic confirmation stage using more precise methods such as NAAT or drug-susceptibility testing. Diagnostic pathways in programmatic settings vary by availability, clinical judgement, and operational constraints, creating systematic differences in TB detection that cannot be reliably adjusted for; and b) we could not verify the quality of CXR training or the accuracy of X-ray interpretation across sites, and many CXRs were read by Medical Officers without specialised training in TB radiology, which may have led to missed abnormalities. **Fourth**, part of the study period overlapped with the COVID-19 pandemic, during which diagnostic service disruptions and mobility restrictions may have affected both screening participation and follow-up. Nonetheless, the large sample size, multi-state coverage, and consistency of findings across sensitivity analyses strengthen the internal validity of the results and support the robustness of the conclusions

## Data Availability & Materials

The data used in this study are routinely collected programmatic data, which has been anonymized for the purposes of research. While the full analytical data can be made available upon reasonable request to the corresponding author, a sample dataset, along with analysis code, is available publicly at GitHub: https://github.com/ridhimasodhi/CXR-Study-JEET-2.0

